# Evaluating the impact of two next generation long-lasting insecticidal nets on malaria incidence in Uganda: an interrupted time series analysis using routine health facility data

**DOI:** 10.1101/2024.10.03.24314858

**Authors:** Adrienne Epstein, Samuel Gonahasa, Jane Frances Namuganga, Martha Nassali, Catherine Maiteki-Sebuguzi, Isaiah Nabende, Katherine Snyman, Joaniter I Nankabirwa, Jimmy Opigo, Martin J Donnelly, Sarah G Staedke, Moses R Kamya, Grant Dorsey

## Abstract

**Introduction:** Malaria remains a significant public health challenge globally, particularly in sub-Saharan Africa, where progress has stalled in recent years. Long-lasting insecticidal nets (LLINs) are a critical preventive tool against malaria. This study investigated the effectiveness of newer-generation LLINs following a universal coverage campaign in Uganda.

**Methods:** Health facility data collected 36 months prior to LLIN distribution and 24 months after LLIN distribution were utilized from 64 sites that took part in a cluster randomized trial comparing two newer-generation LLINs (pyrethroid-PBO and pyrethroid-pyriproxyfen). Using an interrupted time series approach, we compared observed malaria incidence with counterfactual scenarios if no LLINs were distributed adjusting for precipitation, vegetation, seasonality, and care-seeking behavior. Analyses were also stratified by LLIN type and study-site level estimates of transmission intensity.

**Results:** Overall, malaria incidence decreased from 827 cases per 1,000 person-years in the pre-distribution period to 538 per 1,000 person-years in the post-distribution period. Interrupted time series analyses estimated a 23% reduction in malaria incidence (IRR = 0.77, 95% CI 0.65-0.91) in the first 12 months following distribution relative to what would be expected had no distribution occurred, which was not sustained in the 13-24 month post-distribution period (IRR = 0.97, 95% CI 0.75-1.28). Findings were similar when stratified by LLIN type. In the first 12 months following distribution, LLIN effectiveness was greater in the high transmission sites (IRR = 0.67, 95% CI 0.54-0.86) compared to the medium (IRR = 0.74, 95% CI 0.59-0.92) and low transmission sites (IRR = 0.87, 95% CI 0.56-1.32).

**Conclusion:** This study demonstrated a modest reduction in malaria incidence following the distribution of newer-generation LLINs that was sustained for only 12 months, highlighting the need for improved strategies to maintain net effectiveness. Adjusting the frequency of universal coverage campaigns based on local malaria transmission intensity may enhance control efforts.

## BACKGROUND

Major efforts towards malaria control in sub-Saharan Africa have been met with success, resulting in a 44% reduction in malaria incidence from 2000 to 2019 (1, 2). Much of this success has been attributed to the scale-up of long-lasting insecticidal nets (LLINs). Access to LLINs in sub-Saharan Africa has increased markedly in the past two decades, from 5% of households with at least one net in 2000 to 70% in 2022 (2), with many countries in sub-Saharan Africa now distributing LLINs free-of-change in universal coverage campaigns (UCC), typically conducted every three years. Recently, however, progress toward reducing malaria burden has stalled and even reversed course in some high-burden African countries (2). Waning effectiveness of LLINs due to the spread of pyrethroid resistance, changing vector behaviors, poor net adherence, and net attrition are likely contributing to this recent reversal in progress (3–6). Widespread resistance to pyrethroid insecticides has led to the development and distribution of newer generation nets, including those that combine pyrethroids with piperonyl butoxide (PBO), a pyrethroid synergist, or with different insecticides such as pyriproxyfen, an insect growth inhibitor.

Cluster randomized controlled trials (CRT) are considered the optimal method for comparing the efficacy and effectiveness of different LLINs and shaping policy recommendations. While there is ample evidence that newer generation nets are more effective than traditional pyrethroid LLINs (7), less epidemiological evidence of the real-world longitudinal impact of newer generation nets on malaria burden (e.g., cases averted over time) is available. Such evidence is essential for understanding the dynamics of malaria after LLINs are distributed, deciding on the duration between LLIN distribution campaigns, and estimating the cost effectiveness of LLINs. The most rigorous method for quantifying the impact of LLIN distribution would be a CRT including an arm without LLIN distribution. However, such a study design would be unethical given the known benefits of LLINs, and therefore alternative study designs and analytical strategies are needed.

Uganda is one of the high malaria burden countries in sub-Saharan Africa where progress has reversed in recent years. Coverage of LLINs in Uganda is the highest globally (8) due to repeated UCCs conducted approximately every 3 years by the Ministry of Health (MoH) since 2013. Nevertheless, malaria burden remains high and between 2018 and 2022, reported malaria cases increased by 1.7 million from 10.9 million to 12.6 million (2). Widespread resistance to pyrethroid insecticides across Uganda has led to the distribution of newer generation LLINs, including pyrethroid-PBO and pyrethroid-pyriproxyfen LLINs. A CRT was embedded into the 2020-21 UCC to compare these two newer generation nets across 64 sites. In this trial, there were no significant differences between the two LLINs on the incidence of malaria among community members of all ages, or parasite prevalence among children 2-10 years of age, over 24 months following LLIN distribution (9). However, these results did not include estimates of the overall impact of LLIN distribution on malaria incidence over time. By leveraging interrupted time series (ITS) methodologies (10, 11), we used up to 36 months of data prior to LLIN distribution to estimate a counterfactual trend of malaria incidence over 24 months if LLINs had not been distributed. We then compared observed malaria incidence to counterfactual incidence to generate effect estimates for the impact of LLINs over the 24 months post-distribution, aiming to improve our understanding of the real-world effectiveness of newer generation LLINs on malaria burden over time.

## METHODS

### Data source

This study leveraged data from a network of health facility surveillance sites established in 2006 through collaboration between the MoH/National Malaria Control Division and Uganda Malaria Surveillance Program (UMSP) (12). UMSP operates within selected level III/IV health facilities across Uganda, referred to as Malaria Reference Centers (MRCs). At each MRC, individual-level patient data are entered into an electronic database using a standardized register form. Patient information includes demographics (age, sex, and village/parish of residence), whether malaria was suspected, malaria laboratory testing results (either rapid diagnostic test [RDT] or microscopy), diagnoses, and treatments prescribed. UMSP supports health facilities to ensure high quality data, including training, supervision, and adequate stocks of laboratory supplies.

This study used data from 64 MRCs included in a cluster-randomized trial assessing the impact of two newer-generation LLINs on malaria incidence (9). We included 36 months of data pre-2020-21 LLIN distribution (baseline) and 24 months of data post-LLIN distribution; if a site had less than 36 months of baseline data available, we included the maximum amount available (see Supplemental Table 1 for the number of months contributed by each site). Given the variable contribution of each site to the baseline period and the fact that nets were also distributed in 2017-18, we conducted a sensitivity analysis to determine whether limiting the baseline period to 24 and 12 months pre-LLIN distribution impacted the results.

### Study setting and long-lasting insecticide treated net distribution

Details of the parent cluster-randomized trial have been previously reported (9). Briefly, in 2020-21, the Ugandan MoH implemented a UCC, distributing LLINs free-of-charge across the country. As part of this campaign, a cluster randomized trial (LLINEUP2) designed to evaluate the impact of two different LLINs, pyrethroid-PBO LLINs (PermaNet 3.0) and pyrethroid-pyriproxyfen LLINs (Royal Guard), was carried out. Thirty-two districts with high malaria burden not receiving indoor residual spraying (IRS) and selected by the Uganda National Malaria Control Division to receive pyrethroid-PBO LLINs were included in the trial (Figure 1).

**Figure 1.**
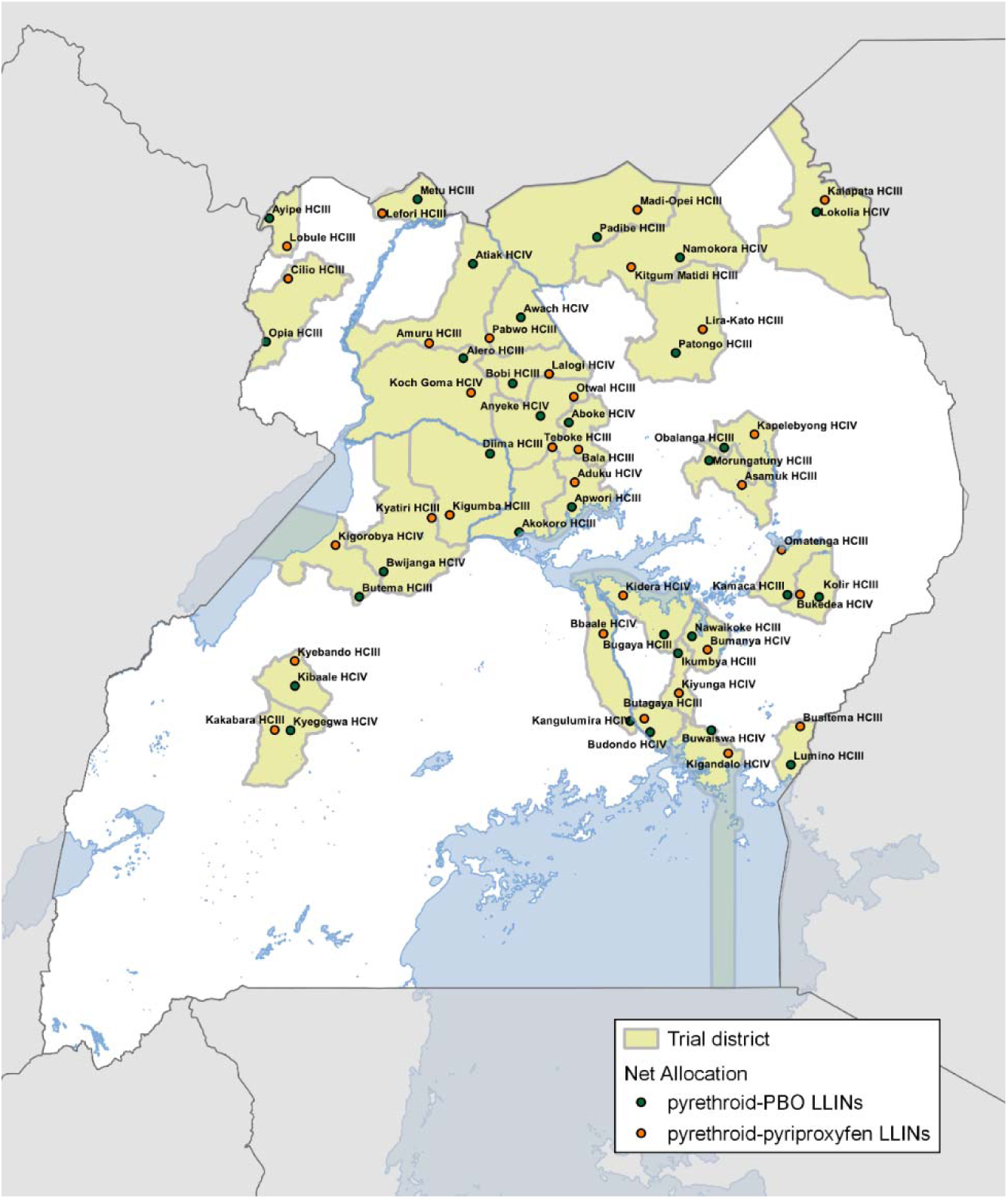
Map of 64 Malaria Reference Centers and their net allocations.

A total of 64 clusters located within these 32 districts (2 per district) were randomized to receive either pyrethroid-PBO or pyrethroid-pyriproxyfen LLINs. A “fried egg” approach was used to measure the impact of the LLINs within the clusters, with the “white” defined as sub-counties receiving LLINs, and the “yolk” as geographically smaller, pre-specified target areas around MRCs, where outcomes were measured. LLINs were delivered to these sub-counties by the Ugandan MoH and partners, adhering to this randomization scheme.

### Measures

The outcome measure for this analysis was monthly malaria incidence in MRC target areas (13). Target areas were defined as a group of 1 or more villages around each MRC, based on the assumption that most patients living within this area with malaria would seek care at the MRC. To validate this assumption, we conducted cross-sectional surveys in randomly selected households from November 2021-March 2022. Of those that were treated for malaria in the last 6 months, 81% went to the MRC. Villages were included if they met the following criteria: (1) did not contain another public health facility, (2) were in the same subcounty as the MRC, and (3) had similar malaria incidence to the village where the MRC is located. Populations of the MRC target areas were determined during enumeration surveys conducted 12 months after the LLIN distribution. The numerator for monthly incidence estimates within target areas was defined as the monthly count of laboratory-confirmed malaria cases among patients residing in the target area (adjusted for target area residents with suspected malaria who did not undergo laboratory testing, and for patients with confirmed malaria whose village of residence was unknown). The denominator was defined as the population of the target areas estimated during enumeration surveys, with a constant growth factor of 0.29% per month (14).

We adjusted for time-varying variables that impact malaria burden and case detection. These include target area-level monthly precipitation lagged by 1 month (15), enhanced vegetation index (16), an indicator variable for calendar month (to account for seasonality), and a monthly count of patients not suspected of having malaria visiting the MRC from the target area (to adjust for care-seeking behaviors over time).

### Statistical analysis

An interrupted time series (ITS) segmented regression approach was taken to estimate the impact of the LLIN distribution on malaria incidence over a 24-month period. The following segmented regression model was estimated:

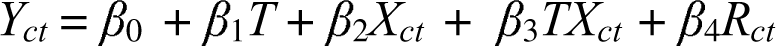

where *Y_ct_* is the outcome (malaria incidence) in cluster *c* at time *t*, *T* is the time elapsed since the start of the study in months, *X_ct_* is a dummy variable indicating the pre-LLIN distribution period (0) or post-intervention period (1) for cluster *c* at time *t*, and *R_ct_* is the vector of covariates for cluster *c* at time *t*. *β*_0_ represents the baseline outcome level at the start of the study (t=0), *β*_1_ represents the change in outcome associated with a one-month increase in the pre-LLIN period, *β*_2_ represents the level change in the outcome after the LLIN distribution, and *β*_3_ represents the additional change in the slope after the LLIN distribution. Poisson regression using a generalized estimating equation was used to model the count of malaria cases in cluster *c* at month *t*, with an offset of the logged population denominator. We included an autoregressive order of 1 correlation structure to account for autocorrelation over time at the cluster level. The resulting ITS model was used to estimate the counterfactual (unobserved) trend of malaria incidence in the absence of the LLIN distribution for each month by setting *X_ct_* to zero. Incidence rate ratios were calculated by comparing the observed incidence to the counterfactual incidence, with bootstrapped 95% confidence intervals.

Our primary analysis estimated the impact of the LLIN distribution pooled across study arms. A secondary analysis allowed the slope change to differ by LLIN arm by including a 3-way interaction term to determine whether the impact of the LLIN distribution differed by net type. We conducted an additional analysis with a 3-way interaction term including a categorical variable for baseline incidence to estimate whether the impact of the LLINs differed across transmission intensities. Baseline incidence was defined by dividing the sites into quartiles and categorizing them into low (100 to 412 per 1,000 person-years [PY]), medium (412 to 765 per 1,000 PY), and high (765 to 2440 per 1,000 PY, the upper two quartiles).

## RESULTS

Across the 64 sites included in the analysis, a total of 3,565,639 outpatient visits were recorded over the study period; 1,505,974 in the pre-LLIN distribution period and 2,059,665 in the post-LLIN distribution period (Table 1). Of these visits, 822,835 were observed within the pyrethroid-PBO arm and 683,139 within the pyrethroid-pyriproxyfen arm in the pre-distribution period and 997,735 and 1,061,930, respectively, in the post-distribution period. Malaria incidence within target areas averaged 827 cases per 1,000 PY in the pre-distribution period and 538 per 1,000 PY in the post-distribution period across all sites. Pre-distribution incidence was 769 cases per 1,000 PY in the pyrethroid-PBO arm and 896 per 1,000 PY in the pyrethroid-pyriproxyfen arm; these figures declined to 501 per 1,000 PY and 576 per 1,000 PY in the post-distribution period, respectively. A total of 16 sites were classified low transmission, 16 sites as medium transmission, and 32 as high transmission. In low transmission sites, malaria incidence in target areas averaged 434 per 1,000 PY in the pre-distribution period and 270 per 1,000 PY in the post-distribution period. These figures were 774 per 1,000 PY and 556 per 1,000 PY in medium transmission sites and 1,261 per 1,000 PY and 778 per 1,000 PY in high transmission sites, respectively.

**Table 1.**
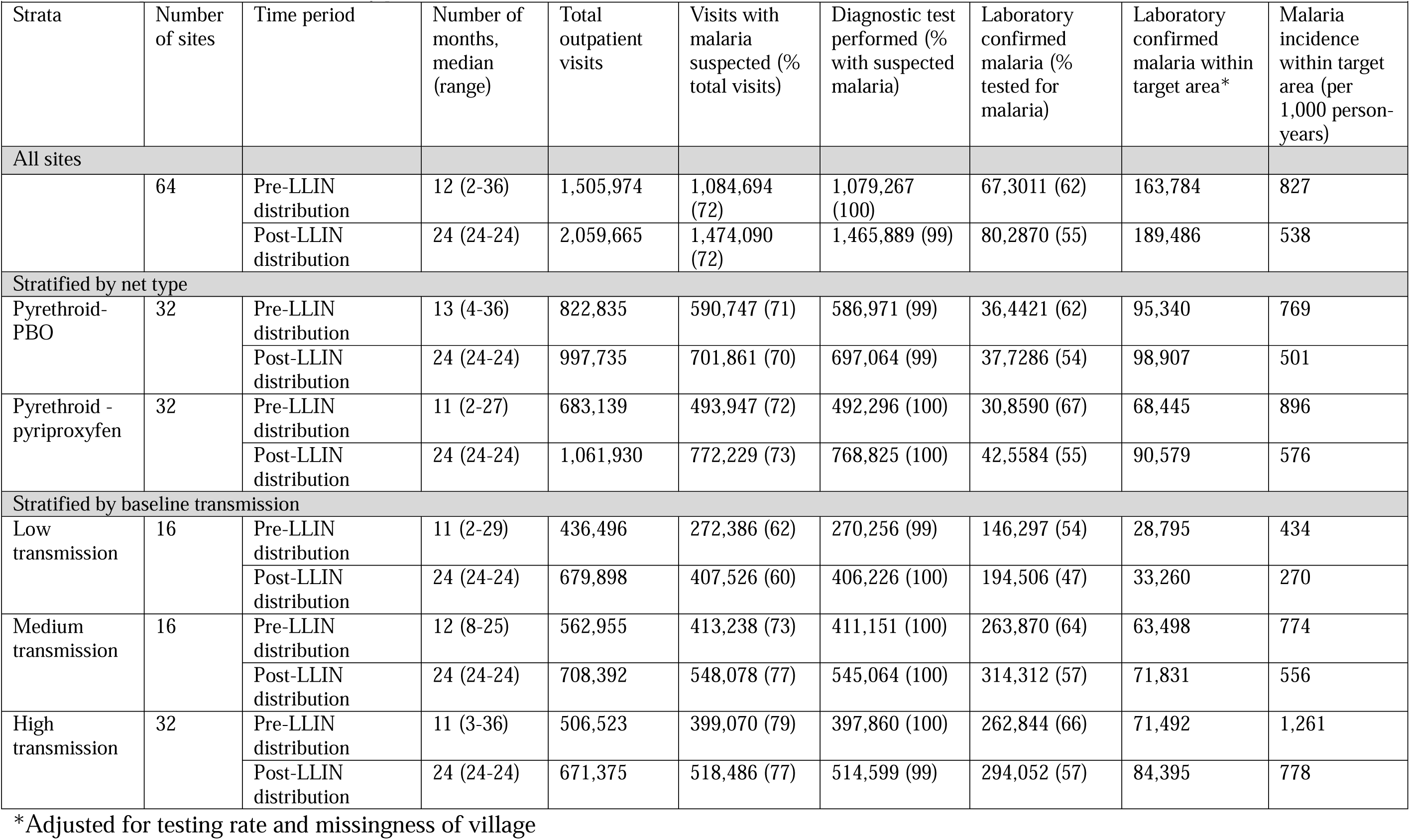
Descriptive statistics over the study period.

Results from the ITS analysis comparing observed and counterfactual malaria incidence pooled across all sites in the 24 months after the LLIN distribution are show in Figure 2 and Table 2. In the first 12 months after the distribution, observed malaria incidence was 23% lower than counterfactual incidence under the conditions of no LLIN distribution (IRR = 0.77, 95% CI 0.65-0.91, Table 2). In months 13-24 post-distribution, observed malaria incidence was 3% lower than the counterfactual, but we could not rule out a null or positive association (IRR = 0.97, 95% CI 0.75-1.28). These results were unchanged when varying the baseline period to 12 and 24 months (Supplemental Figure 1 and Supplemental Figure 2).

**Figure 2.**
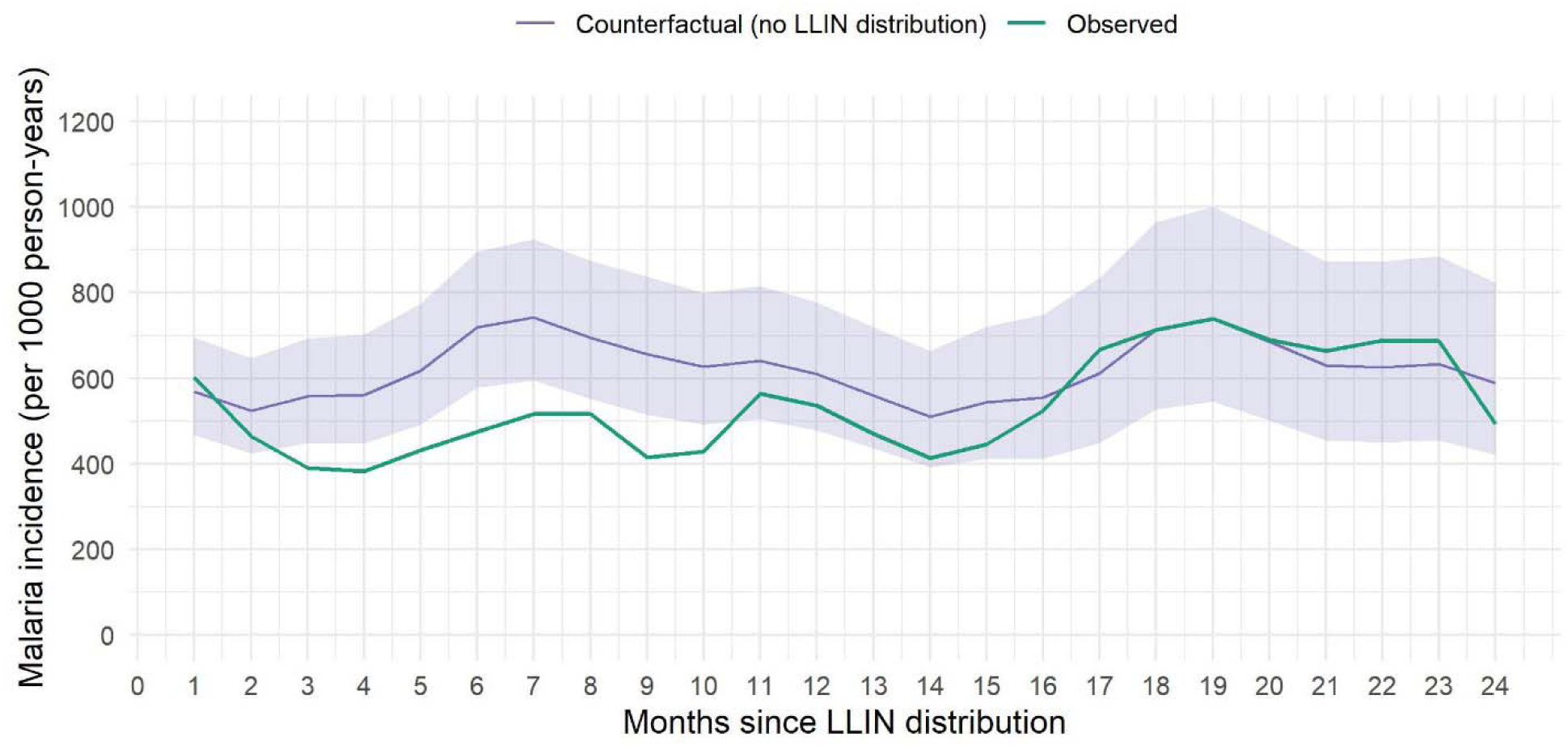
Observed and modeled counterfactual monthly malaria incidence over the 24 months after the LLIN distribution.

**Table 2.** Incidence rate ratios comparing observed malaria incidence to counterfactual malaria incidence modeled using interrupted time series methods.

|  |  |  |  |
| --- | --- | --- | --- |
|  | Pre-LLIN distribution | Months 1-12 post-LLIN distribution | Months 13-24 post-LLIN distribution |
| Strata | IRR (95% CI) | IRR (95% CI) | IRR (95% CI) |
| All sites |  |  |  |
|  | 1.06 (0.89, 1.34) | 0.77 (0.65, 0.91) | 0.97 (0.75, 1.28) |
| Stratified by net type |  |  |  |
| Pyrethroid-PBO | 0.91 (0.79, 1.08) | 0.71 (0.58, 0.88) | 1.01 (0.74, 1.53) |
| Pyrethroid-pyriproxyfen | 1.52 (0.97, 2.19) | 0.78 (0.62, 0.98) | 0.81 (0.54, 1.53) |
| Stratified by baseline transmission |  |  |  |
| Low transmission | 1.32 (0.89, 1.75) | 0.87 (0.56, 1.32) | 1.69 (0.76, 4.08) |
| Medium transmission | 0.98 (0.85, 1.15) | 0.74 (0.59, 0.92) | 0.93 (0.65, 1.46) |
| High transmission | 1.22 (0.93, 1.57) | 0.67 (0.54, 0.86) | 0.73 (0.50, 1.12) |

Results stratified by LLIN type are show in Figure 3 and Table 2. We detected no difference in the post-LLIN distribution change in slope between net types (3-way p-value = 0.18). In the first 12 months after the LLIN distribution, malaria incidence was 29% lower (IRR = 0.71, 95% CI 0.58-0.88) in the pyrethroid-PBO arm and 22% lower (IRR = 0.78 95% CI 0.62-0.98) in the pyrethroid-pyriproxyfen arm comparing observed to counterfactual incidence. From months 13-24 post-distribution, these effect estimates were attenuated and no significant difference was observed for the pyrethroid-PBO arm (IRR = 1.01 95% CI 0.74-1.53) nor for the pyrethroid-pyriproxyfen arm (IRR = 0.81, 95% 0.54-1.53).

**Figure 3.**
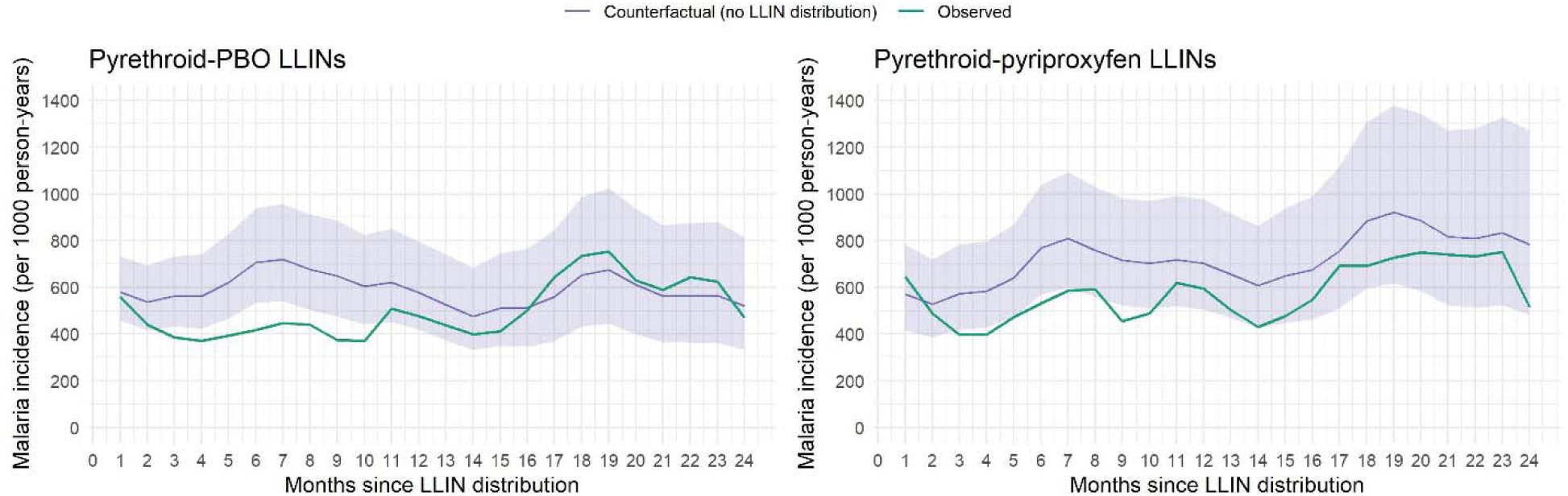
Observed and modeled counterfactual monthly malaria incidence over the 24 months after the LLIN distribution, stratified by net type.

Results stratified by transmission intensity are shown in Figure 4 and Table 2. We detected a significant difference in the post-LLIN distribution change in slope between net types (joint 3-way p-value = 0.0004). In the first 12 months post-distribution, we did not observe a significant difference in malaria incidence in low transmission sites (IRR = 0.87, 95% CI 0.56-1.32). We did, however, observe a 26% reduction in medium transmission sites (IRR = 0.74, 95% CI 0.59-0.92), and a 33% reduction in high transmission sites (IRR = 0.67, 95% CI 0.54-0.86). In months 13-24 post-distribution, we did not observe a significant difference between observed and counterfactual malaria incidence in low transmission sites (IRR = 1.69, 95% CI 0.76-4.08), medium transmission sites (IRR = 0.93, 95% CI 0.65-1.46), nor in high transmission sites (IRR = 0.73, 95% CI 0.50-1.12), though we continued to observe a gradient of effectiveness by transmission intensity similar to that observed in the first year.

**Figure 4.**
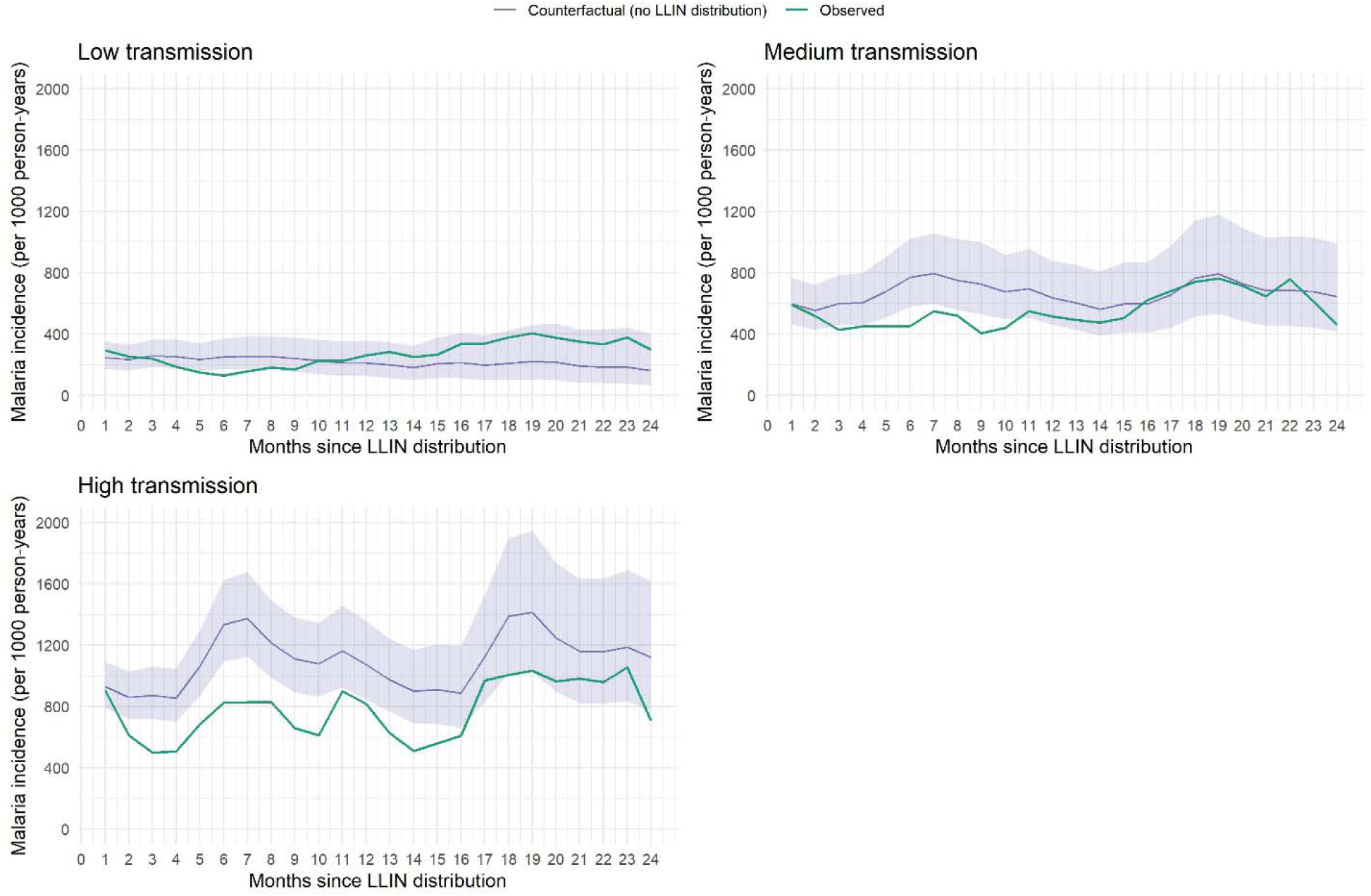
Observed and modeled counterfactual monthly malaria incidence over the 24 months after the LLIN distribution, stratified by baseline malaria incidence.

## DISCUSSION

In this analysis, we utilized community-level malaria incidence data from a CRT to estimate the longitudinal impact of a LLIN UCC using newer-generation nets in Uganda. Our findings suggest that, in this setting, LLINs reduced malaria incidence by 23% in the first year after the UCC, after which effectiveness waned to undetectable levels through the second year post-distribution. These findings were consistent across the two newer-generation net types distributed (pyrethroid-PBO and pyrethroid-pyriproxyfen), with similar effect sizes.

These findings point to a relatively modest and short-lived effectiveness of LLINs in this setting compared to what is generally expected of nets. The present study estimated the effectiveness of LLINs distributed in a “real world” setting, which may account for its more modest effect estimates compared to rigorously conducted CRTs that included control arms (i.e. no bed nets) (17). However, such “placebo-controlled CRTs” were conducted decades ago and the true effectiveness of bed nets may have waned over time. Other observational studies aimed at capturing LLIN effectiveness in “real world” settings have found similar results to this study. In observational studies comparing malaria burden pre-and post-UCC in Burundi and Madagascar, LLINs were associated with modest declines in malaria in the first year post-UCC, but these declines were no longer detectable in years 2-3 post-distribution (18, 19). In Rwanda, PBO nets continued to be effective in the second year post-UCC, but standard nets did not (20). In Malawi, a UCC of PBO and standard pyrethroid-treated nets was associated with reductions in malaria incidence in only the first malaria season post-distribution, but not the second (21).

Multiple factors may have contributed to the relatively modest impact and waning effectiveness of LLINs observed a year after LLIN distribution. First, the number of LLINs distributed during the UCC may have been inadequate, especially as this campaign was carried out during the early years of the COVID-19 pandemic. A 2023 analysis found that household coverage of the 2020-21 UCC was high (94.1%) (22). However, although immediate post-distribution data was not available at all the sites, cross-sectional surveys were conducted at 12 of the sites 1-4 months post-distribution as part of a separate study and revealed that only 60% of household reported adequate LLIN ownership (defined as at least one LLIN per 2 household members) (23). Second, post-distribution coverage studies in many settings have found that retention and use of nets are imperfect and reduce over time following distribution (24–26). Indeed, findings from cross-sectional surveys conducted 12- and 24-months after the LLINEUP2 distribution found that adequate LLIN ownership dropped from 58% to 40%, and use (self-report of household residents sleeping under an LLIN the previous night) fell from 75% to 63%. Third, physical net integrity also degrades over time (24–29). Findings from a trial nested within the previous 2017-18 Ugandan UCC that included pyrethroid-PBO LLINs found that nets experienced an average 80% increase in holed area from 12 to 25-months post-distribution (30). An additional trial in Tanzania found that the functional survivorship of pyrethroid-pyriproxyfen nets was 1.9 years, with only 8.6% in serviceable condition by 36 months (31). While holes found on nets may not markedly reduce the community effect of LLINs against mosquito populations, they may reduce personal protection from bites (32). Furthermore, research suggests that perception of physical integrity is a primary driver for household net retention (33). Fourth, reductions in net bioefficacy likely contributed to the observed waning. In previous cluster randomized trials in Uganda (30) and Kenya (34), pyrethroid-PBO LLINs experienced steep reductions in bioefficacy in the second and third years, respectively, after distribution. While pyrethroid-pyriproxyfen LLINs have demonstrated high bioefficacy in laboratory and experimental hut studies (35), little is known about the longitudinal bioefficacy of these nets in ‘real world’ settings; trials in Tanzania and Benin are ongoing (36, 37). Disentangling the primary causes of declining LLIN effectiveness after a UCC is essential for designing interventions that improve their longevity. A 2020 modeling study that included data from seven trials of both conventional and pyrethroid-PBO nets found that non-use had a larger effect on LLIN impact compared to physical or chemical integrity (6). More research is needed, including qualitative studies, to better understand factors contributing to changes in LLIN effectiveness over time.

We found no difference in the longitudinal impact of LLINs by net type. These findings reaffirm the results of the LLINEUP2 trial, which found no significant difference in 24-month malaria incidence (9). We did, however, find that LLIN effectiveness differed by baseline transmission intensity, such that nets had greater impact in areas with higher baseline malaria incidence, with some additional evidence that effectiveness lasted longer in these areas. This finding echoes observational cohort data from sites with differing transmission intensity in Uganda (38). These results are unsurprising, given that LLINs are likely to have the greatest impact in areas with more malaria-infected mosquitoes. In Uganda, LLIN distributions are conducted on a country level. The findings from this study suggest that the location and frequency of future UCCs could vary depending on malaria burden in the administrative unit. Additional studies, including those focused on cost-effectiveness of LLIN distributions by transmission level, could help to inform this.

A key strength of this study is its outcome measure, malaria incidence measured continuously with high quality data from local health facilities. We leveraged a network of established enhanced health facility-based surveillance sites embedded in public health facilities across Uganda. Health facility data can be challenging to work with due to high rates of clinical diagnosis and aggregate reporting (2), but its utility lies in its wide geographic spread and longitudinal nature. The Uganda Malaria Surveillance Program acts as an intervention in these public health facilities, improving data quality and case management: data are reported at the patient level, diagnostic testing rates are markedly high, and missingness of variables is near-zero. Furthermore, by capturing data on where patients reside and identifying and enumerating target areas around the health facilities, we can continuously and robustly measure malaria incidence on a large scale, at a high temporal resolution, and for a relatively low cost, allowing for longitudinal measurement of the impact of interventions, including this LLIN UCC. This approach can be used in the future to estimate the impact of future UCCs or other interventions, including indoor residual spraying.

This study is not without limitations. Importantly, due to resource constraints, we did not have data from the LLINEUP2 trial on physical integrity, chemical composition, nor bioefficacy; we could therefore not examine the potential contribution of each factor toward LLIN impact. In addition, we had varying amounts of baseline data from different MRCs, with as low as 2 months of baseline data contributing to the model for one MRC, which introduces uncertainty into our counterfactual model. Finally, the ITS model may be impacted by unmeasured confounding. For example, if changes in care-seeking due to the COVID-19 pandemic impacted changes in malaria incidence captured at health facilities, our results may be flawed. However, an analysis assessing the potential impact of the first year of the COVID-19 pandemic at Malaria Reference Centers found no impact of the pandemic on malaria cases and non-malarial visits at health facilities (39).

## CONCLUSIONS

In most high burden countries in sub-Saharan Africa, UCCs are typically conducted every 3 years. There is mounting evidence from observational studies, however, that this timing is too infrequent given degradation in net retention, use, physical integrity, and bioefficacy. This study further contributes to this literature, with longitudinal data on malaria incidence suggesting that distributed newer-generation LLINs were effective for only 12 months after the distribution. Future work aimed at identifying factors that contribute to reductions in LLIN effectiveness is essential for designing interventions aimed at enhancing their longevity. Furthermore, international donors and National Malaria Control Programs may consider reducing the spacing between UCCs, particularly in areas with higher malaria burden, and bolstering continuous distributions through alternative avenues ensure net effectiveness remains high.

## Data Availability

The datasets generated and/or analyzed during the current study will be available at ClinEpiDB (https://clinepidb.org).

## Acknowledgements

We would like to thank the administration and staff at the Infectious Diseases Research Collaboration, and the HMIS officers and in-charges at each of the Malaria Reference Centers.

## Ethics approval

This study involves human participants but this study is exempt from ethical review; all data are routinely collected and deidentified.

## Patient and public involvement

Patients and/or the public were not involved in the design, or conduct, or reporting, or dissemination plans of this research.

## Author note

The reflexivity statement for this paper is available in Appendix S1.

## Contributions

AE conceived of the study with input from GD. SG, JFN, MJN, KS and IN collected the data, with oversight from SGS and MRK. IN and GD managed the data. AE designed and conducted the data analysis. AE drafted the manuscript with guidance from GD, SGS, MJD, and SG. All authors reviewed the manuscript, provided input, and approved the final version for publication.

